# Development and Validation of an Intrinsic Capacity Score in the UK Biobank Study

**DOI:** 10.1101/2024.01.09.24301076

**Authors:** Melkamu Bedimo Beyene, Renuka Visvanathan, Muktar Ahmed, Beben Benyamin, John R. Beard, Azmeraw T. Amare

## Abstract

**Background:** In 2015, the World Health Organization introduced the concept of intrinsic capacity (IC) to define the individual-level characteristics that enable an older person to be and do the things they value. This study developed an IC score for UK Biobank (UKB) study participants and validated its use as a tool for health outcome prediction, understanding healthy aging trajectories, and genetic research.

**Methods:** Our analysis included data from 45,208 UKB participants who had a complete record of the ten variables included in the analysis. Factor adequacy was tested using Kaiser–Meyer– Olkin (KMO), Barthelt’s, and the determinant of matrix tests, and the number of factors was determined by the parallel analysis method. Exploratory and confirmatory factor analyses were employed to determine the structure and dimensionality of indicators. Finally, the IC score was generated, and its construct and predictive validities as well as reliability were assessed.

**Results:** The factor analysis identified a multidimensional construct comprising, one general factor (IC) and five specific factors (locomotor, vitality, cognitive, psychological, and sensory). The bifactor structure showed a better fit (comparative fit index = 0.995, Tucker Lewis index = 0.976, root mean square error of approximation = 0.025, root mean square residual = 0.009) than the conventional five-factor structure. The IC score, generated using the bifactor confirmatory factor analysis has good construct validity, as demonstrated by an inverse association with age (lower IC in older age; beta = -0.035 (95%CI: -0.036, -0.034)), frailty (lower IC score in prefrail, beta = -0.104 (95%CI: (-0.114, -0.094) and frail, beta = -0.227 (95%CI: -0.267, -0.186) than the robust), and Charlson’s comorbidity index (CCI) for incident cases (a lower IC score associated with increased CCI, beta, = -0.019 (95%CI: -0.022, -0.015). The IC score also predicted incident CCI (a one-unit increase in baseline IC score led to lower CCI, beta = 0.147 (95%CI: -0.173, - 0.121)).

**Conclusion:** The bifactor structure showed a better fit in all goodness of fit tests. The IC construct has strong structural, construct, and predictive validities and it is a promising tool for monitoring aging trajectories.

**Highlights:**

- Large biobank studies should be leveraged with intrinsic capacity scores developed.
- Factor analysis confirmed IC as one general factor and five specific factors.
- Better goodness of fit observed with the bifactor model versus conventional structure.
- The bifactor model IC score has a good construct and predictive validity.
- The IC score can be used as a single reliable measure of IC.

## Introduction

In 2015, the General Assembly of the United Nations adopted the 2030 Agenda for Sustainable Development. Of the 17 sustainable development goals, the first was to ensure that all human beings are able to fulfill their potential. In the same year, the World Health Organisation (WHO) introduced the concept of intrinsic capacity (IC) to guide our understanding of the realization of that potential [1]. The conceptual roots of IC can be traced to the writings of the philosopher Martha Nussbaum [2] who defined health and wellbeing in terms of people’s capacities, i.e. their opportunity to do and be what they value. The concept has now found use in medicine, health, social, and behavioral science as a lens through which to anticipate and understand human behavior in all its manifestations[3–9].

WHO experts defined IC as the composite of all the physical and mental capacities of the individual. It has since been proposed that this might be operationalized and measured using five critical domains — cognitive, sensory, locomotor, vitality, and psychological [10]. Personal traits in these domains are genetically determined[11–28]. However, over the life course, these are modified by complex and dynamic age-related biological changes which themselves are influenced by a range of environmental stimuli. Peak values are attained in early adulthood, but start to decline with increasing age[3, 29].

Since the introduction of IC, extensive research has been conducted to measure and validate this construct, as well as to assess its relevance in medical, social, and behavioral sciences [3–5, 7–9]. These studies have established a relationship between IC and various factors such as biological biomarkers [30–32], lifestyle[33–36], socio-economic status[3, 31, 37], health outcomes[3–6], and functional abilities [3, 4, 38, 39]. However, many of these studies rely on relatively small sample cohorts, and comprehensive IC constructs are emerging in large global biobanks such as the UK Biobank, where extensive measurements of diverse biological and clinical characteristics are accessible to facilitate further investigations into the longitudinal health implications of IC changes. This research gap underscores the critical need for developing and integrating IC constructs in large-scale biobank studies and as well as validating their potential to predict longer-term health outcomes such as frailty, falls, and quality of life over time.

Beyond its proposed applications in promoting health and well-being, IC scores have potential use to better understand the biological underpinnings of healthy aging, including the interplay of genetic and environmental factors contributing to age-related health decline, as well as the origins of various diseases. For example, by identifying specific genetic factors associated with IC, researchers can gain a deeper understanding of the complex interplay between genetics, lifestyle, and aging, paving the way for more personalized and targeted interventions for older individuals.

The aim of this study was to develop and validate an approach to estimating IC in one of the world’s largest longitudinal repositories including genetic markers, the UK biobank (N = 500,000). By generating an IC score using data from the UK Biobank, this research lays the groundwork for future investigations into the genetic basis of IC and its complex relationship with aging processes.

## Methods

### Study population

The UK biobank is a large-scale, multicenter prospective cohort study that recruited participants aged 40-69 years, living in Scotland, England, and Wales [40, 41]. The baseline assessment was conducted between 2006-2010. Using a self-completed touch-screen questionnaire and brief computer-assisted interviews, several characteristics related to demographic, socioeconomics, environmental, lifestyle choices and health parameters have been collected. Moreover, comprehensive physical and functional assessments have been conducted, along with the collection of biological samples such as blood, urine, and saliva for subsequent biobanking and biochemical analyses. Further details on the measurements, study design, and data collection are available in the UK biobank online protocol (http://www.ukbiobank.ac.uk). After the exclusion of participants with missing data for variables relevant to IC measurement, the final analysis was conducted on a sample of 45,208 individuals.

### Steps of IC index development and validation

Building on the structure originally proposed by the WHO [7], we implemented a stepwise process to develop the IC index (Figure 1). First, an extensive literature search was conducted to identify variables that represent and measure the five critical IC domains, namely cognitive, sensory, locomotor, vitality, and psychological capacities. Next, each of the identified variables was cross-checked for their availability in the UKB dataset, ensuring a reasonable sample size. Using this approach, we selected ten variables relevant to our study, including working memory, fluid intelligence, duration of moderate activity, duration of walking, anxiety, exhaustion, Haemoglobin concentration, forced expiratory volume in one second, hand grip strength and hearing difficulty. The detailed descriptions of how these variables were measured are provided in supplementary table 1.

**Figure 1.**
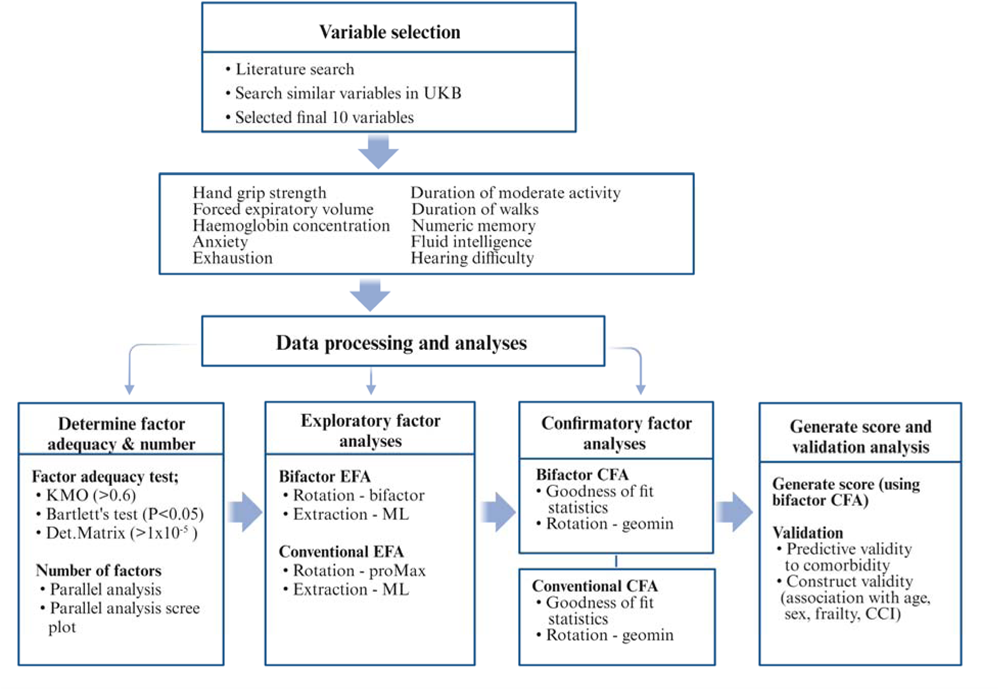
Flow chart showing steps of variable selection, data management, IC index development, and validation. **UKB:** UK biobank**, EFA:** exploratory factor analysis**, CFA:** Confirmatory factor analysis**, Det.Matrix:** Determinant of matrix**, ML:** maximum likelihood**, CCI:** Charlson’s comorbidity index.

Third, factor analysis techniques were implemented, and IC scores were generated using the CFA within the structural equation modeling framework. Finally, we evaluated the construct validity of IC score by assessing its association with age, sex, and frailty and also examined the predictive validity of the IC score by testing its capacity to predict incident Charlson’s comorbidity index (CCI). Frailty was measured according to Fried frailty phenotype (FP) method [42]. Indicators for measuring frailty in the UK biobank were established in previous studies such as Hanlon et al., 2018 [43]. Details of how this variable was measured are provided in supplementary methods 1. The CCI was computed using standard statistical methods suggested by Jonas F. Ludvigsson and colleagues [44] that were based on ICD-10 disease codes for 17 diseases [Myocardial infarction, Congestive heart failure, Peripheral vascular disease, Cerebrovascular disease, Chronic obstructive pulmonary disease, Chronic other pulmonary diseases, Rheumatic disease, Dementia, Hemiplegia, Diabetes without chronic complication, Diabetes with chronic complication, Renal disease, Mild liver disease, liver special, Moderate-severe liver disease, Peptic ulcer disease, Malignancy, Metastatic cancer, HIV/AIDS]. In this study, the incident CCI score was generated taking into consideration diseases that were diagnosed in the period up until 31 December 2022 and after enrolment in the study.

### Statistical Analyses

In this study, we performed (a) EFA (both conventional and bifactor EFA) to examine the structure and dimensionality of selected variables, (b) CFA, within the framework of structural equation modeling (SEM), to test the goodness of fit of the structure and dimensionality identified in the EFAs, and subsequently compute IC composite scores, (c) regression analysis to validate the IC scores.

The first step in the factor analysis was an assessment of factor adequacy, which included examining the KMO measure, Bartlett’s chi-square test of sphericity, and determinant of matrix tests. A KMO value greater than 0.6, a significant Bartlett’s chi-square test of sphericity (P<0.05), and a determinant value greater than 1x10^-5^ were considered indicative of good factor adequacy [45]. Next, the number of factors was determined using the parallel analysis method and examination of the associated scree plot [46]. Eigenvalues greater than one (the number of scree points above the cut-off line on the parallel analysis scree plot) were counted to determine the number of factors. After testing factorability and determining the number of factors, the conventional EFA and bifactor EFA were applied to explore the structure and dimensionality of the dataset. The conventional EFA was performed using the “psych” package with the “factanal” function, employing the “ProMax” rotation method of an oblique type. For the bifactor EFA, the “bifactor” rotation method was selected. In both cases, the maximum likelihood (ML) method was considered as appropriate for factor extraction [47].

The identified structures and dimensions from EFAs were tested using corresponding CFAs, conventional CFA and Bifactor CFA. The lavaan package was used for the CFA, employing the goemin rotation method. The goodness-of-fit of these models/structures was assessed using various statistical tests, including the Root-Mean-Square Error of Approximation (RMSEA), Comparative Fit Index (CFI), Tucker-Lewis index (TLI), and Root Mean Square Residual (RMSR) that were obtained from both conventional and bifactor CFAs. RMSEA value of <0.06, CFI >= 0.95 TLI >= 0.95, and RMSR < 1.0 were considered as indicators of a good fit [48].

The goodness of fit statistics obtained from both conventional and bifactor CFAs was used for model comparison, with the bifactor CFA ultimately chosen for constructing scores for the general IC factor and the five IC domains (cognitive, locomotive, psychological, vitality, and sensory). To evaluate the reliability of the general IC construct, we computed Omega hierarchical. A higher omega hierarchical value (>0.70) indicates a stronger association between the observed variables and the latent construct, indicating greater reliability[49].

Lastly, the construct and predictive validities of the developed IC score were assessed. Construct validity was determined through linear regression analysis using IC as the outcome variable and age, sex, and frailty as independent variables, whereas predictive validity was examined by assessing how baseline IC predicts incident CCI. Here, the CCI index (outcome variable) was calculated for diseases diagnosed after baseline assessment (incident cases).

All the above statistical analyses were performed using R-Studio version 4.2.2 and STATA V.17 programs.

## Results

In this study, a total of 45,208 individuals, with a mean (SD) age of 55.9 (7.6) years, 54% females, were included. At baseline assessment of participants’ frailty status, the majority were categorized as robust (60.7%) or prefrail (37.9%), while only 584 (1.4%) were classified as frail. According to the morbidity data collected until December 31, 2022, most of the participants (71.1%) have no diagnosis of diseases that were included in the Charlson index calculation (CCI = 0). The maximum CCI recorded was 14 (range 0-14). There were 1013 (2.3%) deaths during the follow-up period (from baseline assessment up until December 31, 2022). The detailed characteristics of the participants can be found in Table 1.

**Table 1.** Description of socio-demographic and outcome variables by intrinsic capacity score quartiles (UKB; n=45,208)

| Variables | Category (n) | Grouped by IC scores quartiles |  |  |  |
| --- | --- | --- | --- | --- | --- |
|  |  | Q1<br>(< -0.44) | Q2<br>(-0.44 to -0.03) | Q3<br>(-0.03 to 0.41) | Q4<br>(≥0.41) |
| Sex | Male (20731) | 2347 (5.3) | 3634 (8.1) | 5638 (12.6) | 8886 (19.9) |
|  | Female (24477) | 8914 (19.9) | 7633 (17.1) | 5423 (12.1) | 2222 (5) |
| Age category<br>(in years) | 40-49 (11817) | 1085 (2.4) | 2449 (5.5) | 3404 (7.6) | 4879 (10.9) |
|  | 50-56 (11698) | 2281 (5.1) | 3108 (7) | 3253 (7.3) | 3056 (6.8) |
|  | 57-61 (11362) | 3566 (8) | 3156 (7.1) | 2584 (5.8) | 2056 (4.6) |
|  | 62-72 (9820) | 4329 (9.7) | 2554 (5.7) | 1820 (4.1) | 1117 (2.5) |
| Frailty | Robust (27168) | 5870 (13.1) | 6765 (15.1) | 7063 (15.8) | 7470 (16.7) |
|  | Prefrail (16945) | 5150 (11.5) | 4348 (9.7) | 3888 (8.7) | 3559 (8) |
|  | Frail (584) | 241 (0.5) | 154 (0.4) | 110 (0.3) | 79 (0.2) |
| Charlson's<br>Comorbidity<br>index | Zero (31749) | 7038 (15.8) | 7894 (17.7) | 8175 (18.3) | 8642 (19.3) |
|  | One (5230) | 1747 (3.9) | 1373 (3.1) | 1135 (2.5) | 975 (2.2) |
|  | Two (4265) | 1295 (2.9) | 1088 (2.4) | 1018 (2.3) | 864 (1.9) |
|  | Over two (3453) | 1181 (2.6) | 912 (2) | 733 (1.6) | 627 (1.4) |
| Number of<br>deaths in the<br>follow-up period | Deceased (1013) | 338 (0.8) | 283 (0.6) | 204 (0.5) | 188 (0.4) |
|  | Alive (43,684) | 10923 (24.4) | 10984 (24.6) | 10857 (24.3) | 10920 (24.4) |

### Development of IC score

Using EFA and CFA, and reliability analyses, we developed and validated the IC score. The results from three tests assessing factor adequacy, including the KMO (measure > 0.6), Bartlett’s test of sphericity (P < 0.05), and the determinant of matrix test (<1x10^-5^), collectively indicated factorability and supported the presence of underlying patterns that can be effectively extracted.

The parallel analysis and examination of the associated scree plot identified five factors (Figure 2). In this parallel analysis scree plot, five scree points were above the cutoff horizontal line indicating that the optimal number of factors that can be retained from the factor analysis is five. These factors were previously labeled by WHO experts as vitality, locomotive, psychological, sensory, and cognitive domains.

**Figure 2.**
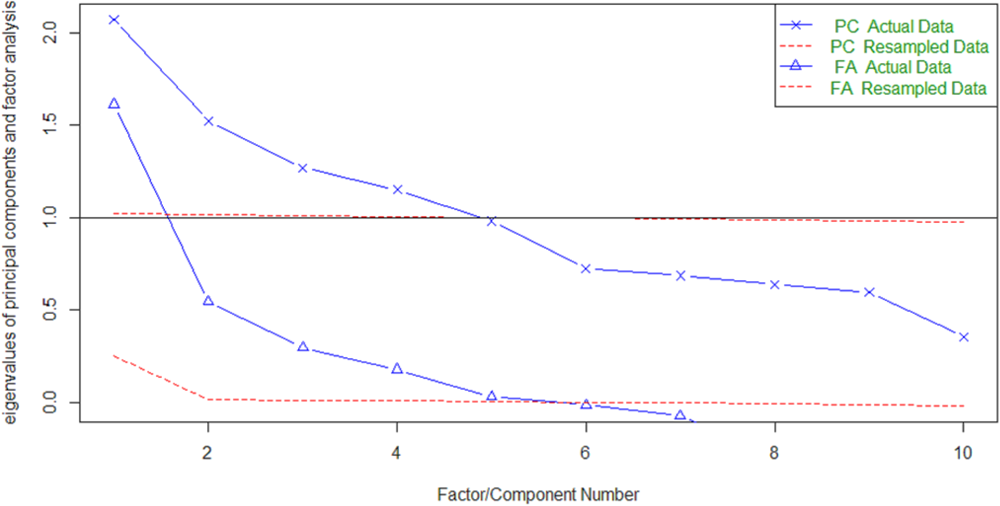
The parallel analysis scree plot where the x-axis represents the number of factors, and the y-axis represents the corresponding eigenvalues. **PC**: Principal component, **FA**: Factor analysis.

The findings from the conventional EFA (Tables 2 and 3) showed a clear loading of the indicators onto five factors with no significant cross-loading. The factor loadings underscored a substantial correlation between the observed indicators and the underlying latent IC construct and its five domains.

**Table 2.** Factor loadings from the conventional exploratory factor analysis (UKB; n=45,208).

|  | **Factor1** | **Factor2** | **Factor3** | **Factor4** | **Factor5** |
| --- | --- | --- | --- | --- | --- |
| Numeric memory |  |  | 0.39 |  |  |
| Fluid intelligence |  |  | 0.75 |  |  |
| Duration of moderate activity |  | 0.95 |  |  |  |
| Duration of walks |  | 0.41 |  |  |  |
| Anxiety |  |  |  | 0.44 |  |
| Exhaustion |  |  |  | 0.66 |  |
| Hemoglobin concentration | 0.33 |  |  |  | 0.42 |
| Forced expiratory volume | 0.78 |  |  |  |  |
| Hand grip strength | 0.78 |  |  |  | 0.16 |
| Hearing difficulty |  |  |  | -0.10 | -0.21 |

**Table 3.** Factor loadings from the bifactor exploratory factor analysis (UKB; n=45,208).

|  | **Vitality** | **IC** | **Cognition** | **Locomotion** | **Psychology** | **Sensory** |
| --- | --- | --- | --- | --- | --- | --- |
| Numeric memory | 0.09 | -0.19 | **0.32** | 0.05 | -0.02 | 0.04 |
| Fluid intelligence | 0.06 | -0.23 | **0.78** | 0.00 | 0.00 | 0.00 |
| Duration of moderate activity | 0.01 | 0.61 | 0.01 | **0.79** | 0.00 | 0.04 |
| Duration of walks | 0.01 | 0.48 | 0.00 | **0.09** | 0.02 | **0.61** |
| Anxiety | -0.08 | -0.02 | -0.06 | 0.02 | **0.45** | 0.01 |
| Exhaustion | -0.08 | -0.03 | 0.01 | -0.01 | **0.63** | 0.01 |
| Hemoglobin concentration | **0.57** | 0.18 | 0.03 | -0.14 | -0.04 | -0.16 |
| Forced expiratory volume | **0.73** | -0.18 | 0.04 | 0.07 | 0.01 | 0.09 |
| Hand grip strength | **0.85** | 0.00 | 0.02 | 0.00 | -0.04 | -0.01 |
| Hearing difficulty | -0.05 | -0.13 | -0.01 | 0.07 | -0.10 | **0.09** |

**IC:** Intrinsic capacity; labeled in bold in the table indicate the corresponding domain that each indicator loads to.

The CFA tested the structure and dimensionality identified by the corresponding EFA, indicating a satisfactory fit across all goodness of fit statistics, with the bifactor structure a better fit than the five-factor structure. Specifically, for the conventional CFA, the model fit statistics were CFI = 0.977, TLI = 0.959, RMSEA = 0.033 (95%CI: 0.031 - 0.034), and RMSR = 0.019. On the other hand, for the bifactor CFA model, the goodness of fit values were CFI = 0.995, TLI = 0.976, RMSEA = 0.025 (95%CI: 0.023, 0.028, and RMSR = 0.009. The final bifactor CFA model revealed that the indicators were loaded into one general IC factor and five distinct latent factors that represented the five IC domains (Figure 3). For instance, the variable grip strength had a factor loading of 0.58 on the general factor (IC) and a loading of 0.80 on its corresponding specific domain (vitality). Finally, the score of both the general IC and the five IC domains were derived from the bifactor CFA under SEM framework. The IC scores (z-standardized scores) for the general and domain-specific domains follow an approximately normal distribution, except for the locomotion domain, which was slightly skewed to the right. The general IC score ranges from -2.74 to 5.47 (Figure 4 and supplementary Figure 1).

**Figure 3.**
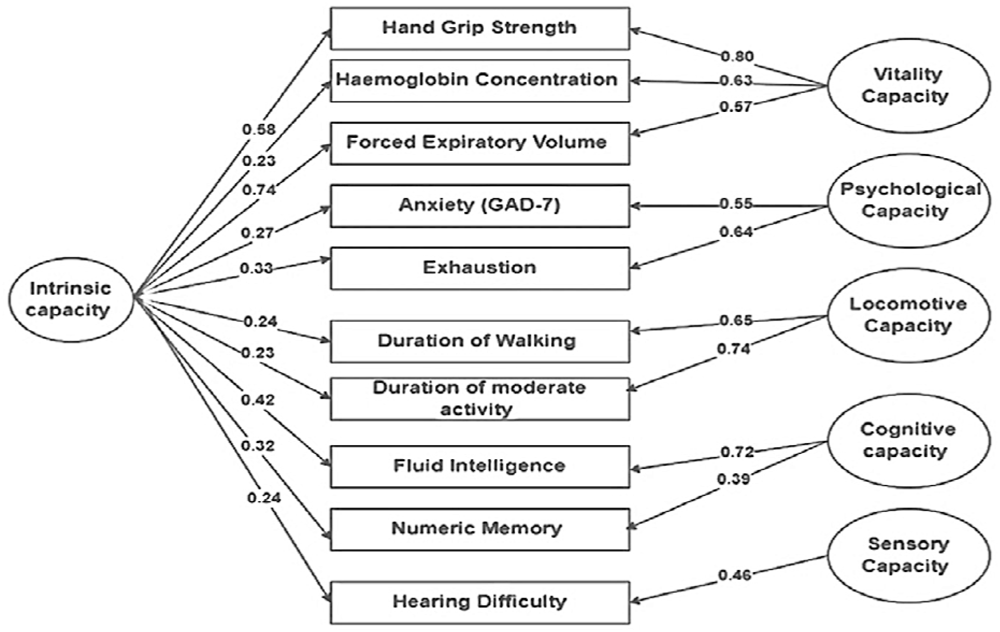
The bifactor model of intrinsic capacity (general construct and the five domains) **GAD:** Generalized anxiety disorder

**Figure 4.**
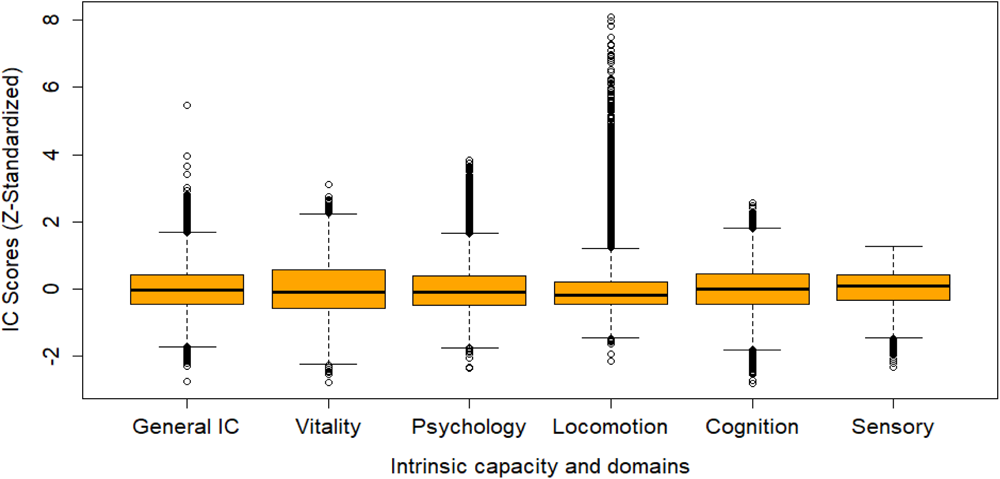
A box plot showing the distribution of Z-standardized IC scores (y-axis) across the IC domains (x-axis). The data presented in the box and whisker plots ranged from the minimum to the maximum IC scores. The lower and upper boundaries of the box correspond to the 25^th^ and 75^th^ percentiles, respectively, and the central line within the boxes represents the median value.

### Validation of the IC Score

Results from the construct and predictive validities, as well as reliability checks, confirmed the validity of the IC score. Findings from the linear regression (construct validity) analysis using the general IC as the outcome variable revealed statistically significant relationships between IC and age, sex, frailty, and CCI for incident cases. Specifically, the results showed that with each year’s increase in age, the IC score decreases by 0.035 units (95%CI: -0.036, -0.034). Men, on average, had a 0.623 higher IC score (95%CI: 0.614, 0.633) compared to women. Participants categorized as prefrail showed, on average, a 0.104 units lower IC score (95%CI: -0.114, -0.094) compared to robust individuals, while frail participants showed, on average, a 0.227 lower IC score (95%CI: -0.267, -0.186) compared to robust ones. Furthermore, the analysis indicated that lower CCI, was associated with higher IC, B = -0.019 (95%CI: -0.022, -0.015) (see Table 4). The scatter plot along with the linear regression line (Figure 5) showed that for both males and females, as age increases on average the IC score decreases consistently. Similarly, Figure 6 shows the finding that non-frail participants have on average higher IC scores than the prefrail. The frail and prefrail participants have lower IC than the non-frail participants (Figure 6)

**Figure 5.**
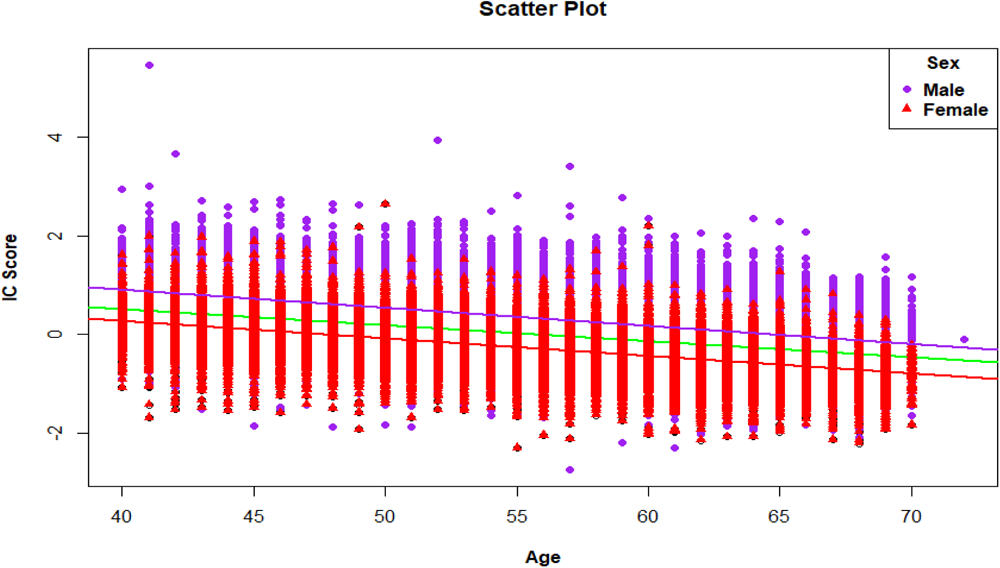
Scatter plot showing the relationship between age and intrinsic capacity stratified by sex (UKB; n=45,208). The predicted regression line, shown in green for the overall sample, pink for the males, and red for the females, illustrates the overall linear relationship between age and IC.

**Figure 6.**
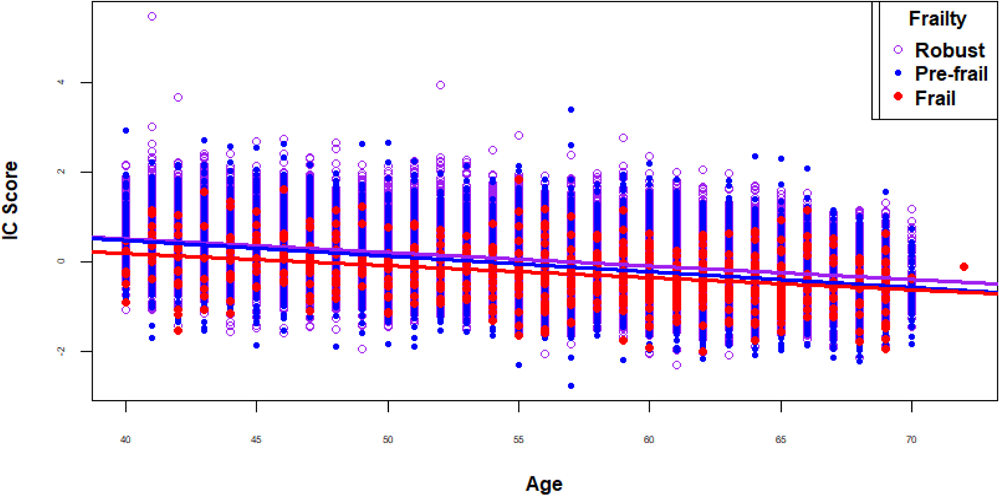
Scatter plot showing the relationship between age and intrinsic capacity stratified by frailty status (UKB; n=45,208). The results from predictive validity analyses demonstrated that the baseline IC significantly predicted the incident CCI. As the baseline IC score increased, the CCI index for the incident cases decreased, and the vice versa (Figure 7). Controlling for the effects of age, sex, and frailty status, a one-unit increased IC score resulted in a 0.147 unit decreased CCI (95%CI: -0.173, - 0.121) (Table 5)

**Figure 7.**
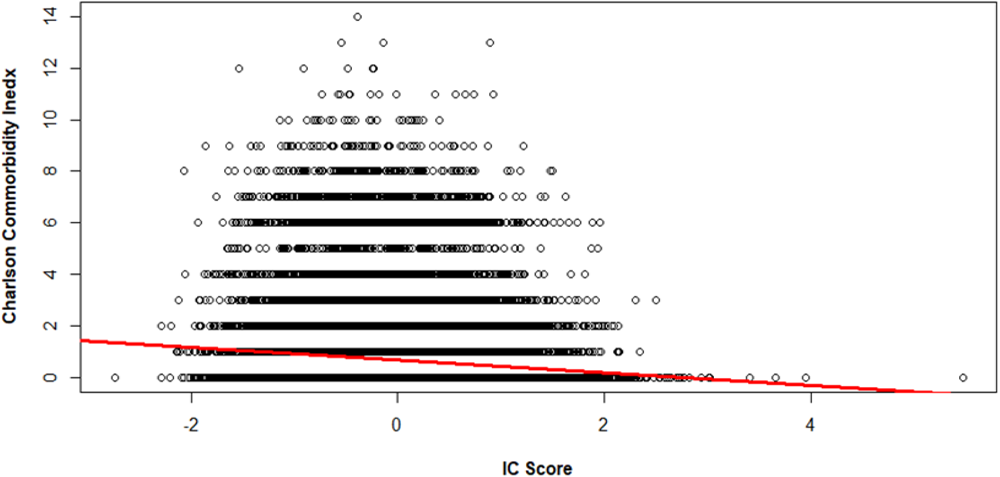
A scatter plot of IC score against CCI with average linear regression line (red); UKB (n=45,208).

**Table 4.** Association of intrinsic capacity with age, sex, frailty, and CCI (crude and adjusted effect estimates and 95%CI)

| Variables | Categories | Beta (95%CI) |  |  |
| --- | --- | --- | --- | --- |
|  |  | Crude | Adjusted |  |
| CCI Score |  | -0.049 (-0.053, -0.045) | -0.019 | (-0.022, -0.015) |
| Age (in years) |  | -0.032 (-0.033, -0.032) | -0.035 | (-0.036, -0.034) |
| **Frailty** | Prefrail vs. Robust | -0.154 (-0.166, -0.141) | -0.104 | (-0.114, -0.094) |
|  | Frail vs. Robust | -0.335 (-0.386, -0.283) | -0.227 | (-0.267, -0.186) |
| Sex | Male vs Female | 0.573 (0.562, 0.583) | 0.623 | (0.614, 0.633) |

**CI:** Confidence interval, **CCI:** Charlson's Comorbidity Index

**Table 5.** The linear relationship between CCI for incident cases and intrinsic capacity (crude and adjusted effect estimates and 95%CI)

| Variables | Categories | Beta (95%CI) |  |
| --- | --- | --- | --- |
|  |  | Crude | Adjusted |
| IC |  | -0.248 (-0.268, -0.227) | -0.147 (-0.173, -0.121) |
| Age |  | 0.038 (0.036, 0.039) | 0.031 (0.029, 0.033) |
| Frailty | Pre frail vs. Robust | 0.210 (0.183, 0.238) | 0.154 (0.127, 0.181) |
|  | Frail vs. Robust | 0.823 (0.706, 0.940) | 0.739 (0.625, 0.854) |
| Sex | Male vs Female | 0.134 (0.107, 0.160) | 0.177 (0.146, 0.208) |

CI: Confidence interval, IC: intrinsic capacity

An omega hierarchical coefficient of 0.96 from the CFA also indicated that the composite IC score was highly reliable and can be used as a single reliable measure of capacity.

## Discussion

The main outcome of this research is the development of a robust IC construct for use within the UK biobank study to enable the subsequent exploration of how genetic markers and gene-environment interactions contribute to the variability of capacity in humans. Five IC domains emerged following EFA, with confirmation using CFA. The bifactor structure had better goodness of fit indices than the correlated factors (conventional) factor structure. Also, the IC score generated using the bifactor structure had good reliability, predictive and construct validities.

In this study, the bifactor structure outperformed the correlated five-factor structure across all computed goodness-of-fit statistics, confirming earlier research [3, 29, 36, 50]. This could be mainly because of the added explanatory power when having a general factor loaded onto the indicators, in addition to specific domains. The concept behind the bifactor structure is such that the general construct (IC) explains variability in the indicators and each of the domains also explains part of the variability in the indicators [51]. Additionally, the high omega hierarchical coefficient (0.96) from the bifactor model suggests that the IC general construct can be used as a single reliable measure of IC.

In the conventional as well as bifactor structures, the vitality domain loads as the first factor. This implies that variations in the vitality domain explain a higher portion of the shared variance than other domains of IC. This could be because it encompasses a wide range of physiological functions and the physiological systems and processes within the body have a substantial impact on an individual’s overall functional abilities and health over time. This finding is consistent with the recent WHO working definition of vitality capacity, which has defined vitality as the underlying physiological determinant of IC, including cardiovascular health, respiratory function, musculoskeletal integrity, metabolic processes, immune function, and other physiological aspects [52].

The IC score generated using the bifactor CFA from SEM has an approximate normal distribution with the z-score ranging from -2.74 to 5.47. The distribution for the domains was approximately normal except for the locomotion domain which had a positively skewed distribution to some extent. This could be because of the sample itself that the UK biobank includes middle-aged adults starting from age 40 years and likely the sample was skewed towards robustness. [53]. The specific questions related to locomotor capacity may also have ceiling effects. The rest of the domains have close to normal distribution with only some slightly outlying observations on either or both sides of the distribution. The sensory domain has outliers only to the negative side (some participants with lower sensory capacity far from others) which implies that sensory capacity, specifically hearing capacity in our case decreases earlier in some people when compared to other domains. Studies show that compared to other functions, sensory functions often exhibit more gradual declines, starting in early adulthood or even before that and becoming more noticeable in later adulthood. However, it’s important to note that the rate and extent of decline can vary among individuals due to other factors too [54, 55]. Consistent with previous research, our analysis revealed a statistically significant relationship between the IC score and age as well as sex. Specifically, it was found that, on average, as age increases, IC tends to decrease, and the vice versa [3, 8, 29, 31, 36, 37, 39, 50, 56–59]. Also, men have on average better IC than women which is also consistent with previous findings [3, 29, 31, 36, 39, 50, 56, 59].

Adding to the growing body of literature on this association [4, 39, 56, 60–64], phenotypic frailty was inversely related (longitudinal and cross-sectional) to IC in this study. Frail participants had on average lower IC than the prefrail and robust ones, and the prefrail participants had lower IC scores than the robust participants. Very recently, a study by Tay et al., 2023 [64] that assessed the association of IC with frailty has found that higher composite IC reduced risk for frailty progression in robust community dwelling adults > 55 years, even after adjusting for age, co-morbidities and social vulnerability. This implies that there is a promise that monitoring for IC decline can help earlier detection of future frailty risk, enabling timely intervention to improve IC that in turn could lead to a delay in the onset of frailty. Opportunity exists within longitudinal studies to identify cut-off scores for IC where risk for future frailty might be increased as this would then allow for interventions to be developed and tested.

Our analysis also demonstrated the potential of IC to predict the likelihood of developing CCI for incident cases. The finding from this analysis showed that there was a statistically significant inverse relationship between the baseline IC and CCI for incident cases in the follow-up. Those participants with lower baseline IC had a higher incident co-morbidity. This finding agrees with evidence from other studies which depict inverse cross-sectional and longitudinal associations between IC and multimorbidity [3, 8, 29, 36, 39, 50, 56, 59]. The relationship with incident CCI is consistent with the “geroscience hypothesis” that age related biological changes increase the risk of most chronic diseases chronic diseases [65]. As with frailty, IC may be an early marker of these changes, identifying those individuals with the highest risk of subsequently developing these conditions.

### Strengths and limitations of the study

One strength of our study lies in the comprehensive assessment of IC. In contrast with previous research on IC within the UK biobank which derived estimates from 4 of 5 domains (missing the cognitive domain) our assessment considers all 5 domains that are generally considered critical components of IC [66]. The downside, however, is that the sample size available for investigation reduced from 443,130 to 45,208, although this remains a large sample size. Another strength of our research was that in contrast to most previous studies of IC which have included older adults only, this research included middle-aged participants (40 to 70 years) thus moving us closer towards a life course approach to IC. Imputation was not used in this study as the number of missing variables was very large and imputation would result in the findings being simulative in nature. It is also important to acknowledge that because the UK biobank Study was not designed as an aging study, important variables that could be included in the IC measurement such as gait speed, self-rated hearing, and vision capacity could not be explored. The UK Biobank may also not represent the entire population due to its volunteer-based recruitment. Participants may be more health-conscious or have specific demographics, potentially leading to selection bias.

### Conclusion and implications

IC is a multidimensional construct, one general domain and five domains emerged from the CFA. The construct generated using the bifactor solution revealed good structural, construct, and predictive validities and is, therefore, suitable for implementation in the UK biobank study to support research relating to IC with that dataset. The conventional correlated five factors solution also showed a satisfactory goodness of fit though the bifactor indices performed better. This research describes an approach as to how IC could be developed and tested using data from existing longitudinal studies. The result of this research, along with a high volume of biological data within the UK biobank, will contribute to building the evidence base towards a better understanding of the genetics and gene-environment interactions underlying IC and healthy aging. The next phase of this research program involves the analysis of longitudinal datasets from global biobanks to understand the biological basis of the interindividual variations in IC and other healthy aging attributes.

## Ethical approval

The UK Biobank has full ethical approval from the NHS National Research Ethics Service (16/NW/0274) and informed consent was obtained before data collection from each participant. This study was conducted using the UK biobank resource under application number 70215.

## Author Contributions

MB Beyene, R Visvanathan, and AT Amare designed the study, secured funding, conducted data analysis, interpreted the results, and drafted the manuscript. M Ahmed provided support on the data analysis. R Visvanathan, and AT Amare supervised the study. All authors (MB Beyene, R Visvanathan, B Benyamin, M Ahmed, JR Beard, AT Amare) critically revised the manuscript for important intellectual content.

## Data Availability

This study was conducted using the UK biobank dataset under application number 70215.

https://www.ukbiobank.ac.uk/enable-your-research/approved-research/investigating-the-genetic-basis-of-human-intrinsic-capacity

## Acknowledgement

We acknowledge the University of Adelaide for the provision of High-Performance Computing facility for data analysis. This study was conducted using the UK biobank resource under application number 70215.

## Data availability

All deidentified data generated or analyzed during this study will be available to approved uses of the UK biobank upon application.

